# Mesocorticolimbic system abnormalities in chronic cluster headache patients: a neural signature?

**DOI:** 10.1101/2021.12.18.21268031

**Authors:** Stefania Ferraro, Jean Paul Medina, Anna Nigri, Luca Giani, Greta Demichelis, Chiara Pinardi, Maria Grazia Bruzzone, Alberto Proietti, Benjamin Becker, Luisa Chiapparini, Massimo Leone

## Abstract

**BACKGROUND:** Converging evidence suggests that anatomical and functional mesocorticolimbic abnormalities support the chronicization of pain disorders.

**METHODS:** We mapped structural and functional alterations of the mesocorticolimbic system in a sample of chronic cluster headache (cCH) patients (n = 28) in comparison to age and sex-matched healthy individuals (n=28) employing structural MRI and resting-state functional MRI (rs-fMRI).

**RESULTS:** Univariate logistic regression models showed that several of the examined structures/areas (i.e., the bilateral nucleus accumbens, ventral diencephalon, hippocampus, and frontal pole, and the right amygdala) differentiated cCH patients from healthy individuals (p<0.05, uncorrected). Specifically, all the significant structures/areas had increased volumes in cCH patients compared to healthy individuals. The examination of the groups suffering from left and right-sided cranial attacks showed a lateralization effect: ipsilateral to the pain ventral diencephalic regions and contralateral to the pain nucleus accumbens discriminated cCH patients from healthy individuals. The rs-fMRI data analyses showed that cCH patients compared to CTRL individuals present robust reduced functional connectivity in the right frontal pole-right amygdala pathway (p<0.05, FDR-corrected).

**CONCLUSION:** Our results showed that cCH patients present anatomical and functional maladaptation of the mesocorticolimbic system, with functional data indicating a possible prefrontal areas’ failure to modulate the mesolimbic structures. These results were opposite to what we hypothesized based on the previous literature on chronic pain conditions. Future studies should assess whether the observed mesocorticolimbic abnormalities are due to the neuroprotective effects of the assumed medications, or to the frequent comorbidity of CH with neuropsychiatric disorders or if they are a genuine neural signature of CH and/or cCH condition.

## 1. Introduction

Cluster headache (CH) is one of the most painful headaches with excruciating unilateral cranial pain, mainly around the orbital and temporal regions, and ipsilateral signs of cranial autonomic activity and/or sense of agitation or restlessness. The episodic form of the disorder is characterized by periods of multiple daily episodes lasting weeks or months followed by attack-free periods. If the symptomatic period does not remit within 12 months, the disorder is considered chronic (cCH) (Olesen, 2018). Although the exact neuropathophysiology of CH is still to be clarified, convergent evidence from clinical, endocrinological, neuroimaging, and animal studies (May et al., 2018; Leone and Bussone, 2009), led to hypothesize a central role of the hypothalamus. In their seminal study, May et al. (1998) observed changes in metabolic activity during CH attacks in a region referred to the posterior hypothalamus (May et al., 1998), supporting the hypothesis that this structure plays a fundamental role in triggering attacks. However, the observation that deep brain stimulation (DBS) of this region and of the ventral tegmental area (VTA) modulate CH attacks within a broad temporal window (weeks or months)(Akram et al., 2016; Leone, 2006) re-framed the abnormal activity of the hypothalamus in a possible deranged functional network (Ferraro et al., 2018; Leone and Bussone, 2009). In our previous work, we showed that cCH patients present dysfunctional connectivity in a network seeded in the ipsilateral-to-the-pain (hereinafter defined as ‘ipsilateral’) posterior hypothalamus and comprising regions of the diencephalic-mesencephalic junction, specifically structures belonging to the midbrain dopaminergic systems, namely the VTA, substantia nigra, and subthalamic nucleus (Ferraro et al., 2018). These observations converge with the evidence of increased plasma levels of dopamine and other related neurotransmitters (D’Andrea et al., 2006, 2017) and with decreased responses of the growth hormone to apomorphine (Lepper et al., 2013) in CH patients, therefore supporting the involvement of the dopaminergic system in CH pathophysiology.

There is now ample evidence that chronic pain disorders are characterized by functional and anatomical abnormalities of the dopaminergic mesocorticolimbic system (Baliki et al., 2012; Vachon-Presseau et al., 2016), a broad network involved in motivational/reward behavior and comprising VTA, nucleus accumbens, amygdala, hippocampus, and prefrontal cortex (Serafini et al., 2020). In chronic pain, this circuit aberrantly responds to salient stimuli (Baliki et al., 2010) and presents altered dopamine transmission (Martikainen et al., 2015). This network seems to play a pivotal role in the transition from acute to chronic pain (Baliki et al., 2012; Vachon-Presseau et al., 2016) such that structural and functional connectivity in the prefrontal cortex-nucleus accumbens network and reduced amygdala and hippocampus volumes predict the development of chronic pain in individuals with subacute back pain. Moreover, when the pain becomes chronic, decreased grey matter volumes are observed in the nucleus accumbens (Baliki et al., 2012). Based on the conceptualization of cCH as a chronic pain disorder, we investigated the presence of structural and functional abnormalities of the mesocorticolimbic system in a relatively large sample of cCH patients (n=28) in comparison to matched healthy individuals (n=28). To this aim, we employed structural magnetic resonance imaging (sMRI) and resting-state functional MRI (rs-fMRI). Specifically, we hypothesized that cCH patients are characterized by decreased volumes of the nucleus accumbens and by aberrant functional connectivity between the nucleus accumbens and prefrontal regions.

## 2. Materials and Methods

### 2.1. Participants

A consecutive sample of 28 patients (5 women; mean age ± SD: 45±11.7 years; age range: 28-70 years) suffering from cCH underwent a cross-sectional MRI study (see table 1 and 2). Two expert neurologists (M.L. and A.P.) made the diagnosis according to the criteria of the International Classification of Headache Disorders (ICHD-3)(Headache Classification Committee of the International Headache Society, 2013). All patients were recruited among the cCH individuals hospitalized to treat the recurrent cluster headache attacks. Patients with a concurrent diagnosis of other primary or secondary headache disorders, cardiovascular diseases, diabetes mellitus, hypertension, or a history of psychiatric conditions were excluded from the study. Except for two, all patients took prophylactic treatment, with 13 patients treated with lithium. A sex and age-matched sample of 28 healthy participants (CTRL), reporting no history of primary headache or chronic pain, was also recruited (mean age ± SD: 45±10.1 years). A subsample of cCH patients (n=19, 2 women; mean age ± SD: 46 ± 10.9 years) and age and sex-matched CTRL individuals (n=18, 2 women; mean age ± SD: 45±10.7 years) also underwent rs-fMRI acquisitions (see table 3). The study was planned and conducted according to the Helsinki Declaration and approved by the Ethical Committee of the IRCCS Neurological Institute Carlo Besta. Each participant gave prior written informed consent.

**Table 1:** Demographic and clinical data of cCH and CTRL individuals submitted to the structural MRI acquisitions. Abbreviations: cCH = chronic cluster headache group, CTRL = control group, NA = not applicable, ys = years, M = mean, SD = standard deviation, left-sided = left-sided CH attacks, right-sided = right-sided CH attacks.

|  | cCH | cCH left-sided | cCH right-sided | CTRL |
| --- | --- | --- | --- | --- |
| # participants | 28 | 12 | 15 | 28 |
| Age (ys; M±SD) | 45±11.7 | 47±22.9 | 48±3.6 | 45±10 |
| Males/Females | 23/5 | 12/2 | 12/3 | 23/5 |
| # daily attacks (M±SD) | 5±2.3 | 4±2.2 | 5±2.5 | NA |
| Disease duration (ys; M±SD) | 7±6.1 | 6± 6.4 | 8±5.9 | NA |

**Table 2:** Current prophylactic treatment in cCH patient. Abbreviations: M = male, F= female, L = left, R = right, BIL = bilateral, DD = disease duration, ys = years.

| Patient ID | Gender | Age (ys) | Side of attack | DD (ys) | # daily attacks | Prophylaxis |
| --- | --- | --- | --- | --- | --- | --- |
| 1 | M | 31-40 | L | 10 | 5 | Verapamil, Valproate |
| 2 | M | 31-40 | L | 3 | 2 | Verapamil, Valproate |
| 3 | M | 61-70 | L | 6 | 3 | Carbolithium, Pizotifen |
| 4 | M | 61-70 | L | 0.33 | 2 | Verapamil, Carbolithium, Dexamethasone |
| 5 | M | 31-40 | L | 2 | 8 | Carbolithium |
| 6 | M | 41-50 | L | 25 | 8 | Verapamil |
| 7 | M | 61-70 | L | 5 | 2 | Verapamil, Carbolithium |
| 8 | F | 31-40 | L | 4 | 2 | Valproate, Verapamil, Carbolithium |
| 9 | M | 41-50 | L | 4 | 4 | Verapamil |
| 10 | M | 41-50 | L | 3 | 5 | Verapamil |
| 11 | F | 51-60 | L | 1 | 2 | Verapamil |
| 12 | M | 41-50 | L | 5 | 2 | Carbolithium |
| 13 | M | 21-30 | R | 7 | 7 | Verapamil, Valproate |
| 14 | M | 31-40 | R | 3 | 8 | Verapamil, Carbolithium |
| 15 | M | 41-50 | R | 2 | 4 | Verapamil, Dexamethasone, Valproate |
| 16 | M | 61-70 | R | 0.5 | 2 | Verapamil, Carbolithium |
| 17 | M | 41-50 | R | 10 | 5 | None |
| 18 | F | 51-60 | R | 1 | 2 | Carbolithium, Dexamethasone |
| 19 | F | 21-30 | R | 2 | 2 | None |
| 20 | M | 31-40 | R | 9 | 6 | Carbolithium |
| 21 | M | 31-40 | R | 18 | 4 | Valproate |
| 22 | M | 41-50 | R | 15 | 8 | Verapamil, Carbolithium |
| 23 | M | 41-50 | R | 9 | 3 | Verapamil, Carbolithium |
| 24 | F | 41-50 | R | 4 | 4 | Duloxetine, Amitripyline |
| 25 | M | 41-50 | R | 10 | 5 | Dexamethasone, Topiramate |
| 26 | M | 41-50 | R | 8 | 4 | Dexamethasone, Verapamil, Carbolithium |
| 27 | M | 51-60 | R | 20 | 2 | Dexamethasone |
| 28 | M | 31-40 | BIL | 1 | 3 | Dexamethasone |

**Table 3:** Demographic and clinical data of the subsamples of cCH and CTRL individuals submitted to the rs-fMRI acquisitions. Abbreviations: cCH = chronic cluster headache group, CTRL = control group, NA = not applicable, ys = years, M = mean, SD = standard deviation, left-sided = left-sided CH attacks, right-sided = right-sided CH attacks.

|  | cCH | cCH left-sided | cCH right-sided | CTRL |
| --- | --- | --- | --- | --- |
| # participants | 19 | 8 | 11 | 18 |
| Age (ys; M±SD) | 46±10.9 | 49±12.9 | 44±9.4 | 44±9.7 |
| Males/Females | 16/3 | 7/1 | 9/2 | 16/2 |
| # daily attacks (M±SD) | 5±2.2 | 5± 2.4 | 5± 2.1 | NA |
| Disease duration (ys;M±SD) | 8± 6.7 | 7± 7.7 | 9± 6.1 | NA |

### 2.2. Descriptive statistics

All statistical analyses were performed using R studio 3.5.1. To test for differences between cCH patients and CTRL group in terms of age, we used the Kruskall-Wallis test. Based on the predominant unilateral cranial symptoms, we investigated possible lateralization of the anatomical mesocorticolimbic alterations examining separately the group reporting left-sided attacks and the group reporting right-sided attacks (see Structural MRI data analyses paragraph). To verify the comparability of these two groups in terms of age, number of attacks per day, and years of chronicization, we employed Kruskall-Wallis tests.

### 2.3. MRI assessments

All participants were scanned with a 3T scanner equipped with a 32-channels coil (Achieva TX, Philips Healthcare BV, Best, NL). The MRI protocol included a volumetric high-resolution structural 3D T1-weighted (T1w) image (TR=9.86ms, TE=4.59 ms, FOV=240×240 mm, voxel size=1 mm^3^, flip angle=8^°^, 185 sagittal slices) and rs-fMRI images (GE-EPI volumes, TR=2800 ms, TE=30 ms, FOV=224×241, voxel size=2.5 mm^3^, flip angle=70^°^, 50 slices with 10% gap acquired in ascending order, 200 volumes). All the patients were scanned at least two hours after the last CH attack.

### 2.4. Structural MRI data analyses

sMRI results of this cohort of patients relative to abnormalities of the telencephalic and cerebellar cortical thickness were already published (Demichelis et al., 2021). For the current sMRI analyses, we employed the data from all the enrolled cCH and CTRL individuals. For each participant, we performed automated segmentation of the cerebral cortical areas and subcortical nuclei (FreeSurfer software v6.0 - http://surfer.nmr.mgh.harvard.edu/) using as input the T1w image. Based on our *a priori* hypothesis, we focused on the following structures/areas of the mesocorticolimbic system: the ventral diencephalon, the hippocampus, the amygdala, the nucleus accumbens, and the medial prefrontal cortex (comprising the medial and lateral orbitofrontal cortex, the frontal pole, and the rostral anterior cingulate cortex). At the end of the automated pipeline (*recon-all* command), to remove pial-white boundary surfaces segmentation inaccuracies, visual inspection and manual correction of the segmentation were conducted by two expert operators (C.P. and G.D.) blind to the condition of the participants (cCH or CTRL). The volume of each structure/area of interest was extracted (from *aseg* and *aparc* files) and expressed as a percentage of total intracranial volume (TIV) to account for inter-subject cerebral volumes variability (Malone et al., 2015).

#### Logistic regression analyses

We assessed the statistical association between each extracted measure across the two groups of participants (cCH and CTRL participants) with respect to cCH diagnosis through univariate logistic regression models, using as covariates age and gender. Specifically, univariate logistic regression models were used to estimate how the volumes of the selected mesocorticolimbic structures affect the probability of belonging to the cCH group. Moreover, we evaluated the same statistical association separately for the left-lateralized attacks group (n=12) and right-lateralized attacks group (n=15). In this case, one patient with recurrent bilateral attacks was excluded from the analyses. We quantified the diagnostic discrimination of the univariate models using the area under a receiver operating characteristics curve (AUC). We used the maximum likelihood estimator to estimate the AUC, providing a score for each patient. To additionally control for possible effects of lithium, we assessed the statistical association between each extracted measure with respect to potential differences between lithium-treated and non-lithium-treated patients at the time of scanning. Moreover, employing the Pearson correlation coefficient, we evaluated whether each investigated structure/area volume in cCH individuals correlated with the years of disease duration and with the number of headache attacks per day. Based on the specific *a priori* hypothesis, we considered results as significant for p<0.05.

### 2.5. rs-fMRI data analyses

CONN toolbox (www.nitrc.org/projects/conn) was employed to analyze the rs-fMRI dataset. rs-fMRI scans underwent a series of standardized preprocessing steps comprising slicetiming correction, realignment, normalization to the MNI152 space, and spatial smoothing (6 mm FWHM Gaussian kernel). Preprocessed data were then denoised according to the CONN default pipeline. We then performed ROI-to-ROI functional connectivity (rs-FC) analysis. The ROIs were extracted from the Harvard-Oxford cortical and subcortical structural atlas, the default CONN atlas. In line with the morphological analyses, we selected the following ROIs: the hippocampus, the amygdala, the nucleus accumbens, and the medial prefrontal cortex (in this atlas identified as the medial frontal cortex, orbital frontal cortex, and frontal pole). The ventral diencephalon ROI was not present in this atlas; therefore, we extracted it from the atlas employed for the structural MRI analyses. In each subject, the mean average time-series of each ROI was computed, and for each possible pair of ROIs, a Fisher-transformed bivariate correlation coefficient was obtained. ROI-to-ROI rs-FC matrices were calculated. One sample t-tests were used to identify significant rs-FC among the investigated regions separately for CTRL and cCH individuals. Using age and gender as covariates, a one-way ANCOVA was employed to analyze between-group differences. Moreover, we investigated the correlation between the ROI-to-ROI rs-FC with the years of the chronicization of the disease and with the number of CH attacks per day in the cCH group. The results were considered significant for p<0.05 FDR-corrected.

## 3. Results

### 3.1. Descriptive statistics

No significant differences emerged between cCH patients and CTRL individuals regarding age. No significant differences were observed between the group of patients with left-sided attacks and the group of right-sided attacks in terms of age, number of daily attacks, and disease duration.

### 3.2. Structural MRI data results

#### Logistic regression analyses

Univariate logistic regression models demonstrated that the bilateral nucleus accumbens, ventral diencephalon, hippocampus, and frontal pole, and the right amygdala significantly discriminated cCH patients from CTRL individuals (AUC range=0.65-0.74; see table 4). Moreover, the odds ratios (OR) of these significant models were positive, thus indicating that increasing the volume of the structure/area increases the likelihood of an individual to be categorized as a cCH patient. Remarkably, larger volumes were observed in the cCH group than in the CTRL group for all the significant structures/areas (table 4). All the models showed that age and gender did not significantly impact the discrimination accuracy; therefore, to avoid a reduction of degrees of freedom, we did not use these covariates in the subsequent morphological analyses. Logistic regression models also showed that the volumes of the right nucleus accumbens, the left ventral diencephalon, the right hippocampus, and the left frontal pole (AUC range=0.65-0.74) discriminated the left-sided attacks cCH group from the CTRL group. Similarly, the volumes of the left nucleus accumbens (AUC=0.79) and the right ventral diencephalon (AUC=0.68) for the right-sided attacks cCH group (see table 4 and figure 1). These results indicated that in both patients groups, the ipsilateral-to-the-pain (hereinafter defined as ‘ipsilateral’) ventral diencephalon and the contralateral-to-the-pain (hereinafter defined as ‘contralateral’) nucleus accumbens discriminated cCH patients from the CTRL individuals. Significantly, logistic regression models of all the investigated structures/areas did not discriminate between lithium-treated patients and non-lithium-treated patients (table 6). In cCH individuals, no significant correlations were observed between the volumes of the investigated structures and the years of the disease duration. A significant inverse correlation was observed between the volumes of the right frontal pole and the number of cluster headache attacks per day (p=0.022; r=-0.43).

**Table 4:** Logistic regression models of the volumes (expressed as a percentage of the total intracranial volume) of the structures/areas of the mesocorticolimbic system estimating the probability of the cCH condition (Overall - 28 cCH individuals), of the left-sided attacks cCH condition (Left-sided attacks -12 cCH patients), and of the right-sided attacks cCH condition (Right-sided attacks -15 cCH patients) in respect to CTRL condition (28 healthy individuals). Abbreviations: OR: Odds Ratio, CI= confidence interval, AUC= area under the curve, R = right, L = left, cCH = chronic cluster headache condition; CTRL = control condition.

| Marker | Overall |  |  |  |  | Left-sided attacks |  |  |  | Right-sided attacks |  |  |  |
| --- | --- | --- | --- | --- | --- | --- | --- | --- | --- | --- | --- | --- | --- |
|  | OR~ | 95%CI | P-value | AUC | 95%CI | OR~ | 95%CI | P-value | AUC | OR~ | 95%CI | P-value | AUC |
| L nucleus accumbens | 4.34 | 1.63-14.54 | 0.008 | 0.73 | 0.59-0.87 | 3.34 | 1.07-12.77 | 0.05 | 0.66 | 6.49 | 1.82-34.25 | 0.01 | 0.79 |
| R nucleus accumbens | 3.44 | 1.18-11.78 | 0.033 | 0.65 | 0.50-0.80 | 4.58 | 1.24-20.72 | 0.03 | 0.67 | 3.81 | 0.92-19.83 | 0.08 | 0.67 |
| L ventral diencephalon | 1.38 | 1.07-1.89 | 0.024 | 0.67 | 0.53-0.81 | 1.64 | 1.13-2.59 | 0.02 | 0.73 | 1.31 | 0.98-1.93 | 0.11 | 0.63 |
| R ventral diencephalon | 1.36 | 1.05-1.90 | 0.040 | 0.65 | 0.50-0.79 | 1.36 | 0.99-2.11 | 0.10 | 0.63 | 1.46 | 1.05-2.18 | 0.04 | 0.68 |
| L hippocampus | 1.23 | 1.01-1.57 | 0.065 | 0.66 | 0.51-0.80 | 1.23 | 0.96-1.64 | 0.11 | 0.66 | 1.25 | 0.98-1.73 | 0.12 | 0.66 |
| R hippocampus | 1.31 | 1.06-1.72 | 0.025 | 0.69 | 0.55-0.83 | 1.50 | 1.10-2.08 | 0.02 | 0.74 | 1.23 | 0.97-1.65 | 0.12 | 0.65 |
| L amygdala | 1.35 | 0.83-2.31 | 0.248 | 0.57 | 0.42-0.73 | 1.73 | 0.87-3.91 | 0.14 | 0.63 | 1.07 | 0.55-2.05 | 0.84 | 0.51 |
| R amygdala | 1.65 | 0.99-2.99 | 0.068 | 0.64 | 0.49-0.79 | 2.07 | 1.00-5.02 | 0.07 | 0.67 | 1.58 | 0.86-3.17 | 0.16 | 0.63 |
| L frontal pole | 1.76 | 1.04-3.21 | 0.047 | 0.65 | 0.50-0.79 | 2.12 | 1.11-4.762 | 0.04 | 0.69 | 1.38 | 0.70-2.86 | 0.36 | 0.59 |
| R frontal pole | 1.81 | 1.06-3.32 | 0.039 | 0.70 | 0.50-0.84 | 1.64 | 0.89-3.26 | 0.13 | 0.70 | 1.76 | 0.96-3.58 | 0.08 | 0.71 |
| L lateralorbitofrontal | 1.02 | 0.93-1.14 | 0.623 | 0.47 | 0.32-0.63 | 1.09 | 0.97-1.26 | 0.17 | 0.43 | 0.95 | 0.80-1.10 | 0.47 | 0.59 |
| R lateralorbitofrontal | 0.98 | 0.88-1.10 | 0.755 | 0.42 | 0.27-0.57 | 1.05 | 0.92-1.20 | 0.46 | 0.51 | 0.85 | 0.67-1.01 | 0.10 | 0.67 |
| L medialorbitofrontal | 1.06 | 0.94-1.22 | 0.369 | 0.54 | 0.38-0.69 | 1.06 | 0.91-1.26 | 0.44 | 0.49 | 1.06 | 0.90-1.27 | 0.50 | 0.55 |
| R medialorbitofrontal | 1.02 | 0.88-1.19 | 0.767 | 0.49 | 0.34-0.65 | 1.13 | 0.94-1.39 | 0.21 | 0.57 | 0.96 | 0.77-1.18 | 0.67 | 0.53 |
| L rostralanteriorcingulate | 0.84 | 0.65-1.05 | 0.141 | 0.37 | 0.22-0.53 | 0.90 | 0.65-1.22 | 0.52 | 0.58 | 0.78 | 0.55-1.05 | 0.12 | 0.65 |
| R rostralanteriorcingulate | 0.89 | 0.70-1.11 | 0.331 | 0.39 | 0.24-0.54 | 0.96 | 0.70-1.23 | 0.74 | 0.62 | 0.74 | 0.51-1.03 | 0.09 | 0.63 |

**Table 5:** Mean and standard deviations of the volumes of the investigated structures/areas (expressed as % of the total intracranial volume - TIV) of the mesocorticolimbic system: Abbreviations: cCH = chronic cluster headache patients, CTRL = control individuals, M = mean, SD = standard deviation, L = left, R = right.

| Marker | cCH |  | CTRL |  |
| --- | --- | --- | --- | --- |
|  | M | SD | M | SD |
| L nucleus accumbens | 0.028 | 0.007 | 0.023 | 0.005 |
| R nucleus accumbens | 0.033 | 0.006 | 0.030 | 0.004 |
| L ventral diencephalon | 0.283 | 0.028 | 0.268 | 0.018 |
| R ventral diencephalon | 0.276 | 0.030 | 0.261 | 0.014 |
| L hippocampus | 0.273 | 0.031 | 0.258 | 0.024 |
| R hippocampus | 0.282 | 0.031 | 0.263 | 0.023 |
| L amygdala | 0.101 | 0.013 | 0.097 | 0.008 |
| R amygdala | 0.116 | 0.012 | 0.110 | 0.009 |
| L frontal pole | 0.061 | 0.011 | 0.055 | 0.010 |
| R frontal pole | 0.071 | 0.009 | 0.065 | 0.011 |
| L lateralorbitofrontal | 0.456 | 0.066 | 0.449 | 0.043 |
| R lateralorbitofrontal | 0.445 | 0.057 | 0.450 | 0.041 |
| L medialorbitofrontal | 0.309 | 0.046 | 0.299 | 0.039 |
| R medialorbitofrontal | 0.319 | 0.041 | 0.316 | 0.031 |
| L rostralanteriorcingulate | 0.116 | 0.028 | 0.122 | 0.021 |
| R rostralanteriorcingulate | 0.141 | 0.026 | 0.151 | 0.021 |

**Table 6:** Logistic regression models of the volumes (expressed as a percentage of the total intracranial volume) of the structures/areas of the mesocorticolimbic system estimating the probability of the lithium-treated cCH condition in respect to the no lithium-treated cCH condition. Abbreviations: cCH = chronic cluster headache, OR = Odds Ratio, CI = confidence interval, AUC = area under the curve, R = right, L = left.

| **Marker** | **OR~** | **95%CI** | **P-value** | **AUC** | **95%CI** |
| --- | --- | --- | --- | --- | --- |
| L nucleus accumbens | 1.31 | 0.47-3.94 | 0.61 | 0.55 | 0.32-0.79 |
| R nucleus accumbens | 1.13 | 0.32-4.08 | 0.85 | 0.52 | 0.30-0.75 |
| L ventral diencephalon | 1.00 | 0.75-1.33 | 0.99 | 0.50 | 0.28-0.73 |
| R ventral diencephalon | 1.10 | 0.85-1.48 | 0.46 | 0.58 | 0.36-0.80 |
| L hippocampus | 0.88 | 0.65-1.13 | 0.36 | 0.43 | 0.21-0.65 |
| R hippocampus | 0.82 | 0.57-1.06 | 0.18 | 0.63 | 0.41-0.84 |
| L amygdala | 0.75 | 0.38-1.35 | 0.36 | 0.46 | 0.23-0.70 |
| R amygdala | 0.62 | 0.28-1.19 | 0.18 | 0.62 | 0.39-0.85 |
| L frontal pole | 0.43 | 0.14-0.96 | 0.07 | 0.68 | 0.47-0.88 |
| R frontal pole | 0.51 | 0.17-1.24 | 0.16 | 0.63 | 0.41-0.85 |
| L lateralorbitofrontal | 0.95 | 0.81-1.07 | 0.45 | 0.549 | 0.33-0.77 |
| R lateralorbitofrontal | 0.89 | 0.71-1.04 | 0.24 | 0.69 | 0.48-0.89 |
| L medialorbitofrontal | 0.95 | 0.78-1.12 | 0.54 | 0.57 | 0.34-0.81 |
| R medialorbitofrontal | 1.01 | 0.83-1.22 | 0.95 | 0.50 | 0.27-0.73 |
| L rostralanteriorcingulate | 1.27 | 0.94-1.80 | 0.14 | 0.62 | 0.39-0.85 |
| R rostralanteriorcingulate | 0.92 | 0.65-1.22 | 0.58 | 0.48 | 0.25-0.70 |

**Figure 1:**
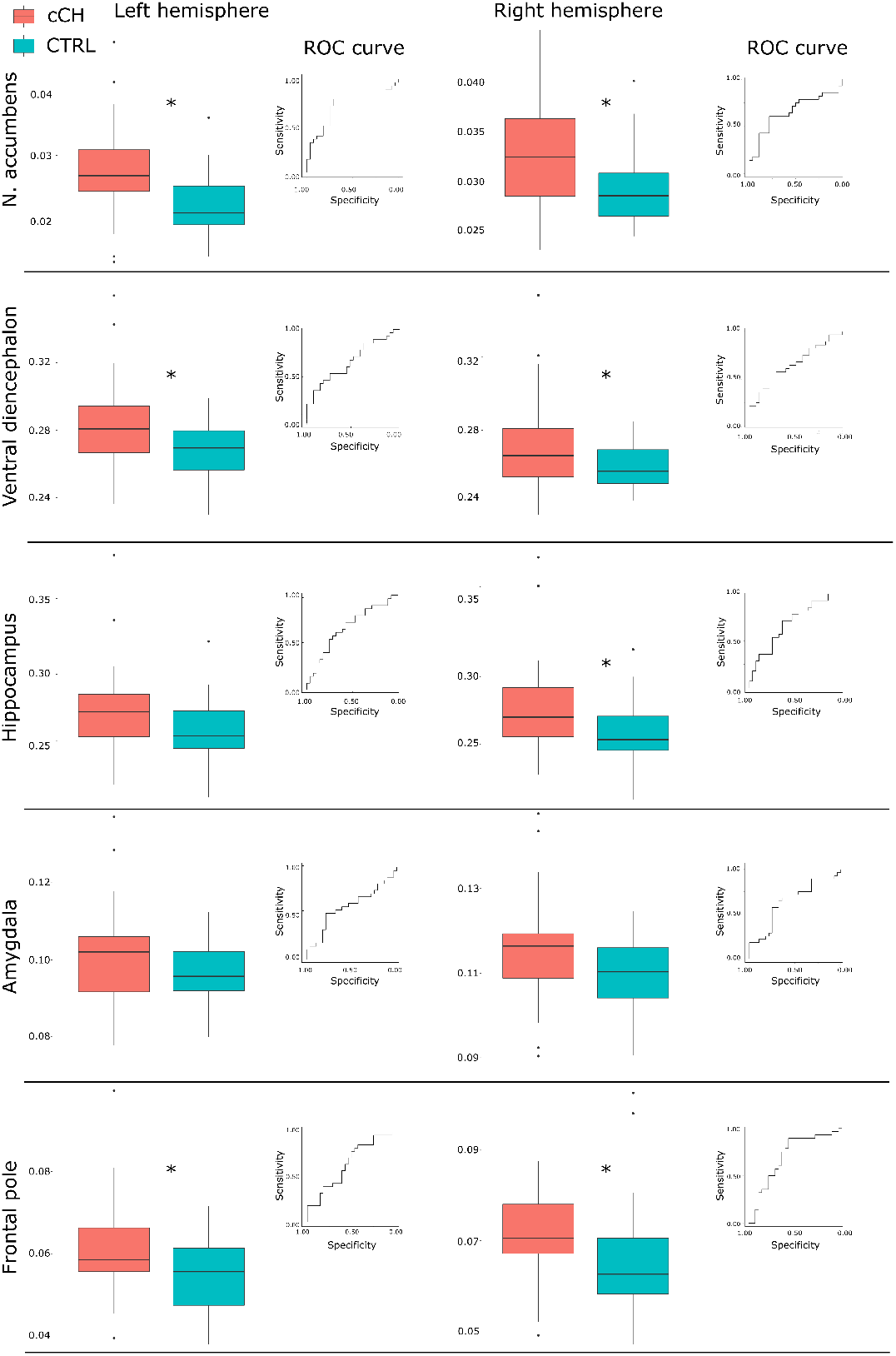
Structural MRI data results: Boxplots of the volumes (expressed as a percentage of the total intracranial volume) of the mesocorticolimbic structures/areas in chronic cluster headache (cCH) and control participants (CTRL) and the respective receiver operating characteristic (ROC) curves for the diagnostic accuracy of the classifiers (univariate logistic regression models). *Structures/areas significantly discriminating cCH patients from CTRL individuals. Abbreviations: cCH = chronic cluster headache patients, CTRL = control individuals.

**Figure 2:**
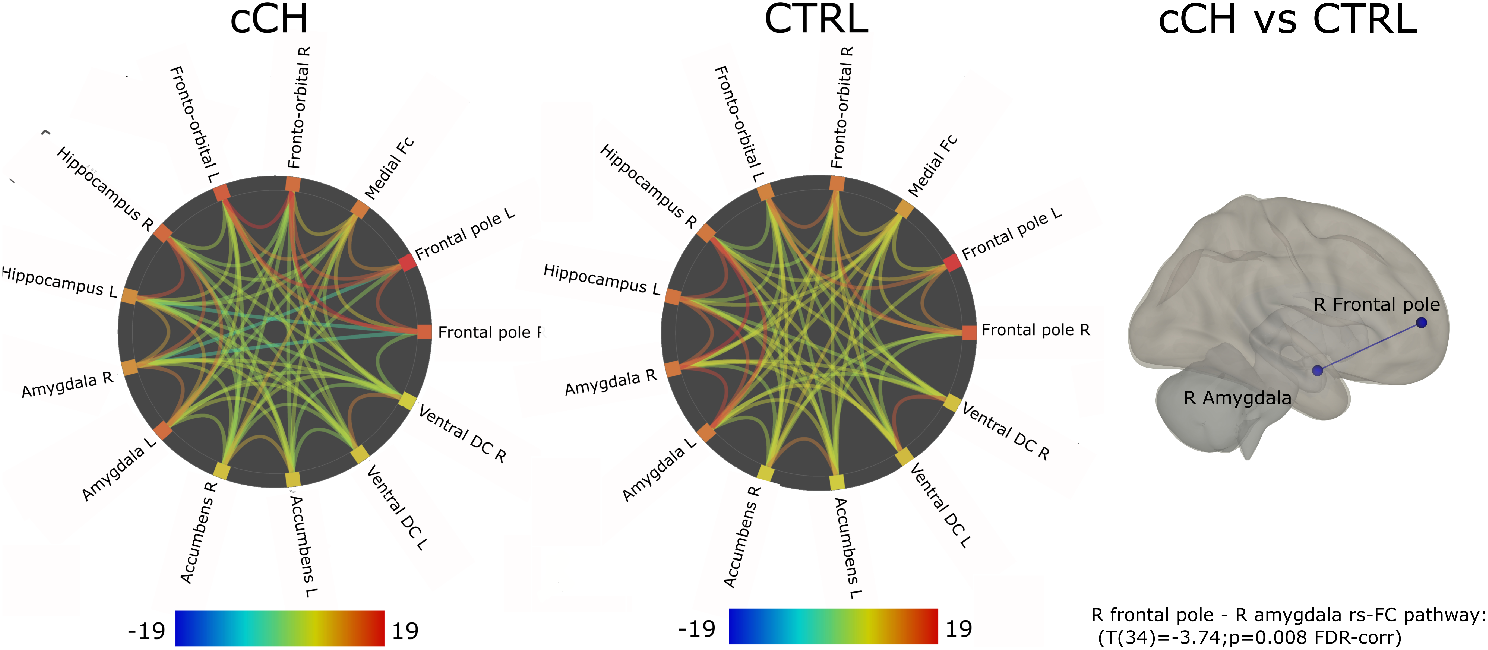
ROI-to-ROI resting-state functional connectivity (rs-FC). Connectome ring representations of the significant ROI-to-ROI rs-FC of the investigated structures/areas of the mesocorticolimbic system for cCH patients, CTRL individuals and representation of the reduction of the rs-FC in the functional pathway right frontal-pole-right amygdala in cCH patients compared to CTRL individuals (p<0.05 FDR-corrected). The color bars represent T values. Abbreviations: cCH = chronic cluster headache patients, CTRL = control individuals.

#### 3.3 rs-fMRI data results

cCH patients compared the CTRL individual showed a robust reduced rs-FC in the right frontal pole-right amygdala pathway (T(34)=-3.74; p= 0.008 FDR-corrected) (see 2). There were no significant correlations between the ROI-to-ROI rs-FC and the years of disease duration, and the number of headache attacks per day.

## 4. Discussion

Based on a strong *a priori* hypothesis, we mapped structural and rs-FC alterations of the mesocorticolimbic system in a relatively large sample of cCH patients compared with an age and sex-matched CTRL group. We report three main robust findings. First, employing logistic regression analyses on sMRI data, we demonstrated that cCH patients can be discriminated from CTRL individuals based on the volumes of several mesocorticolimbic structures/areas, specifically the bilateral ventral diencephalic region, the bilateral nucleus accumbens, the bilateral frontal pole, the bilateral hippocampus, and the right amygdala. Second, the separated investigations of the cCH groups reporting left-sided and right-sided cranial attacks shed light on the lateralization of these abnormalities: cCH patients were discriminated from CTRL individuals by the ipsilateral ventral diencephalic regions and by the contralateral nucleus accumbens. Remarkably, contrary to our hypothesis, all the mesocorticolimbic structures/areas showing significant diagnostic accuracy were larger in cCH individuals than in the CTRL group. Third, employing ROI-to-ROI rs-FC analyses, we showed that cCH patients present reduced rs-FC in the right frontal pole-right amygdala pathway.

There is now accumulating evidence that the mesocorticolimbic system plays a key role in motivated behavior and reward/aversive processing (Kringelbach and Berridge, 2015; Schultz, 2016). More recently, a central role in the transition from acute to chronic pain was also proposed (Baliki et al., 2012; Vachon-Presseau et al., 2016). This broad network originates from the dopaminergic inputs of the VTA targeting several subcortical and cortical brain regions involved in executive, affective and motivational behavior. Key areas of this circuit are also the nucleus accumbens and the medial prefrontal cortex, with a role also for the amygdala and the hippocampus (Navratilova and Porreca, 2014). Noxious stimuli were shown to reduce the activity of most of the mesolimbic dopamine neurons (value-coding neurons), but to increase the activity of a specific subpopulation of dopaminergic neurons (salience-coding neurons), leading to hypothesize the presence of a motivational salience signal rather than an intrinsic value signal of the input (Bromberg-Martin et al., 2010). In agreement, a separate dopaminergic circuit encoding for aversive-predicting stimuli has been identified from the ventral tegmental area to the ventral medial shell of the nucleus accumbens, and excitatory inputs from the lateral hypothalamus to the ventral nucleus accumbens were shown to mediate aversive-related behavior (de Jong et al., 2019). Moreover, neuronal changes in nucleus accumbens were linked to the impairment of motivation in chronic pain (Schwartz et al., 2014). Altogether, the above findings provided converging evidence that maladaptations of the dopaminergic mesocorticolimbic circuit may underlie abnormalities of the affective and motivational mechanisms in chronic pain, in agreement with the long-standing notion that chronic pain conditions are often in comorbidity with affective and emotional disorders (e.g., anxiety, depression) (Bushnell et al., 2013). Based on this background, a series of seminal studies, mainly conducted in back pain, has shown that chronic pain conditions are characterized by abnormal signaling of the end of acute pain in the nucleus accumbens during pain processing (Baliki et al., 2010), abnormalities of the striatal dopaminergic system (Martikainen et al., 2015), and temporal-dependent volume loss of the bilateral nucleus accumbens and insula cortex (Baliki et al., 2012). Moreover, smaller amygdala and hippocampal volumes and increased rs-FC between the prefrontal cortex and nucleus accumbens were shown to be predictive of pain chronification (Baliki et al., 2012; Vachon-Presseau et al., 2016). This last finding, in particular, led to hypothesize the presence of an abnormal emotional signal disturbing the dopaminergic pathway from the ventral tegmental area, which in turn would modulate the thalamocortical network, which would result in an amplification of the nociceptive inputs (Vachon-Presseau et al., 2016). Notably, a recent study on trigeminal neuralgia, a chronic neuropathic pain, found evidence of bilateral volumetric decrease of the nucleus accumbens, ventral diencephalon, and hippocampus, supporting again the the notion of abnormal affective circuits in chronic pain conditions (Hayes et al., 2017).

cCH is a chronic pain condition, therefore the observed anatomical and functional abnormalities in the mesocorticolimbic system were not unexpected. However, instead of reduced volumes as observed in chronic pain, our results showed that cCH patients, compared to CTRL individuals, present increased volumes of most parts of the structures/areas of this system. Very remarkably, we observed that the ipsilateral ventral diencephalon volumes discriminate cCH patients from CTRL individuals, therefore indicating lateralization of the volumes abnormalities, very plausibly linked to the lateralization of the cranial pain. It is very important to note that the observed enlarged ipsilateral ventral diencephalon, a region comprising the hypothalamus, the basal forebrain, the sublenticular extended amygdala (SLEA), and the ventral tegmental area (Makris et al., 2008) is in agreement with the observations of increased volumes of the ipsilateral hypothalamus reported in the seminal study by May et al. (1999) and more recently observed by Arkink et al. (2017) and with the long-standing notion that the hypothalamic/ventral tegmental areas are involved in CH pathophysiology (Akram et al., 2016; Leone and Bussone, 2009; May et al., 2018). Interestingly, the nucleus accumbens presents a contralateral enlargement in cCH patients compared to CTRL participants. Although we have no clear explanation, this result again suggests a lateralization of the morphological abnormalities. It is plausible that in cCH patients, a potential reduced volume of ipsilateral nucleus accumbens, linked to a lateralized effect of this chronic pain condition, is compensated by possible neurotrophic effects of the assumed medications. In contrast, the non-affected contralateral nucleus accumbens might result in increased volumes.

Several and not mutually exclusive explanations may explain the divergent results in the context of the previous studies on chronic pain disorders. First, lithium, a mood-stabilizing medication with a therapeutic effect on cCH patients, is well known for its neurotrophic and neuroprotective effect (Manji et al., 2000). In bipolar disorders, the assumption of lithium is linked to increased volumes of the hippocampus (Yucel et al., 2008), thalamus (Hibar et al., 2016), frontal regions (Abramovic et al., 2016), and with clear effects on the nucleus accumbens shape (Vecchio et al., 2020). The use of lithium might explain the increased mesocorticolimbic volumes, and more specifically, the increased volumes of the contralateral nucleus accumbens, as discussed above. However, the observation that in our sample lithium was assumed, at the time of the MRI examination, in less than half of patients (13 out of 28) and that no significant differences emerged between the mesocorticolimbic volumes of the lithium-treated and non-lithium-treated patients render this possibility less probable. In this regard, we cannot exclude long-term effects of lithium interrupted in the past or that other employed medications in the investigated sample of cCH individuals might provide neuroprotective effects. Other possible explanations for the observed abnormalities include the comorbidity of CH condition with anxiety disorders (Robbins, 2013), linked to enlargement of the nucleus accumbens (Günther et al., 2018), and the use of the licit and illicit substance of abuse by CH individuals (Govare and Leroux, 2014). In this regard, animal studies have consistently shown increased dendritic spines in neurons of the nucleus accumbens in animals treated with morphine (Robinson and Kolb, 1999), cannabis (Kolb et al., 2006), nicotine (Brown and Kolb, 2001), and cocaine (Robinson et al., 2001). A third explanation is that the observed abnormalities of the mesocorticolimbic system could constitute the neural signature of the cCH or CH conditions.

Similarly to the structural results, we observed unexpected findings also for the rs-fMRI data: instead of increased rs-FC in cCH patients between the nucleus accumbens and the prefrontal regions as reported in chronic pain conditions (Baliki et al., 2012; Vachon-Presseau et al., 2016), we observed rs-FC reduction primarily in the right frontal pole-right amygdala pathway. Notably, inhibitory control of the prefrontal cortex over the nucleus accumbens dopamine release was shown to be modulated by the amygdala in animal models (Jackson and Moghaddam, 2001). Therefore, our results might indicate an imbalance of the frontal pole in control over the nucleus accumbens, possibly mediated by the activity of the amygdala. These results support the hypothesis of a failure of the frontal pole – amygdala functional pathway in modulating the mesolimbic activity in cCH patients (Piacentini et al., 2017).

Altogether, these unexpected findings underline the specificity of cCH in respect to other chronic pain conditions. In CH, a chronic pattern may establish since the very beginning of the headache attacks; therefore, factors involved in the pathophysiology must differ from those known in chronic pain and in chronic migraine. This concept is further supported by the clinical experience of cCH cases with sudden and stable remission. Probably relevant to our results, at variance to other chronic pain disorders, is also the paroxysmal nature of CH attacks with sustained pain-free intervals between attacks, different from chronic pain with persistent–continuous pain perception all day long.

Although important for the possible implications, our results should be considered with great caution: we could not control the effects of the assumed medications and of the smoking habits, and we did not assess the impact of anxiety or depression, or other neuropsychiatric disorders in the investigated sample of cCH patients. Moreover, the fact that we did not compute the statistical power before conducting the experiment suggests evaluating the null findings carefully (Colegrave and Ruxton, 2003), especially the results from the sMRI analyses conducted in the subsamples of cCH individuals. Notably, rs-fMRI data were collected in a subsample of cCH patients (n=18), therefore to avoid a further reduction of the statistical power, we did not investigate the lateralization of the ROI-to-ROI rs-FC abnormalities and the impact of the lithium treatment on the ROI-to-ROI rs-FC.

To conclude, we showed that cCH patients present anatomical and functional a maladaptation of the mesocorticolimbic system, however these results were opposite to what we hypothesized based on the previous literature on chronic pain conditions.

Future studies should assess whether the observed anatomical and functional abnormalities in cCH patients are linked to the neuroprotective effects of the assumed medications, or to the frequent comorbidity of CH with neuropsychiatric disorders or if they are a genuine neural signature (as neural bases or epiphenomenon) of CH and/or cCH disorder, promoting the disease itself or the development of the chronicization of the disease.

## Data Availability

All data produced in the present study are available upon request to the authors.

